# Cross-cultural translation and psychometric validation of the French version of the Fear-Avoidance Components Scale (FACS)

**DOI:** 10.1101/2023.07.07.23292355

**Authors:** Arnaud Duport, Sonia Bédard, Catherine Raynauld, Martine Bordeleau, Randy Neblett, Frédéric Balg, Hervé Devanne, Guillaume Léonard

## Abstract

**Background:** The Fear-Avoidance Components Scale (FACS) is a reliable and valid instrument widely used to assess fear-avoidance beliefs related to pain and disability. However, there is a scarcity of validated translations of the FACS in different cultural and linguistic contexts, including the French population. This study aimed to translate and validate the French version of the FACS (FACS-Fr/CF), examining its psychometric properties among French-speaking individuals.

**Methods:** A cross-cultural translation process – including forward translation, backward translation, expert committee review, and pre-testing – was conducted to develop the FACS-Fr/CF. The translated version was administered to a sample of French-speaking adults (n=55) with musculoskeletal conditions. Internal consistency (including confirmatory analyses of the 2 factors identified in the Serbian version), test-retest reliability and convergent validity were then assessed.

**Results:** The FACS-Fr/CF demonstrated high global internal consistency (α=0.94, 95% CI: 0.91-0.96) as well as high internal consistency of the 2 factors identified in the Serbian version (α=0.90, 95% CI: 0.86-0.94 and α=0.90, 95% CI: 0.85-0.94, respectively). Test-retest analysis revealed a moderate (close to high) reliability (ICC=0.89; 95% CI: 0.82-0.94 and r=0.89; *p*<0.005). Convergent validity was supported by significant correlations between the FACS-Fr/CF scores and the Tampa Scale for Kinesiophobia (r=0.82; *p* < 0.005), the Pain Catastrophizing Scale (r=0.72; *p* < 0.005) and the Hospital Anxiety and Depression Scale (r=0.66; *p* < 0.005).

**Conclusion:** The present study provides evidence for the cross-cultural translation and psychometric validation of the FACS-Fr/CF. The FACS-Fr/CF exhibits a high internal consistency, a moderate (close to high) test-retest reliability, and good construct validity, suggesting its utility in assessing fear-avoidance beliefs in the French-speaking population. This validated tool can enhance the assessment and understanding of fear-avoidance behaviors and facilitate cross-cultural research in pain-related studies.

## 1 Introduction

The fear-avoidance (FA) model is proposed as a possible explanation for the transition from acute to chronic pain in some patients [1,2]. According to this model, individuals who perceive their pain as threatening and who catastrophize will tend to develop fear of pain, avoid regular activities, and monitor excessively bodily sensations [1,3,4]. These responses may precipitate physical deconditioning, limit the ability to work and to participate in recreational/familial activities, and foster depression [1,3,4].

Several patient-reported outcome (PRO) questionnaires have been developed to quantify FA related concepts, including the Tampa Scale for Kinesiophobia (TSK) [5], the Pain Anxiety Symptoms Scale (PASS) [6], the Pain Catastrophizing Scale (PCS) [7], and the Fear-Avoidance Beliefs Questionnaire (FABQ) [8]. However, the FA model has significantly evolved in recent years, and none of these questionnaires comprehensively examine all the cognitive, emotional, and behavioral components of the model [9]. Although they have been used in a substantial number of peer-reviewed published research, the psychometric properties of these tools (including construct validity and sensitivity to change) have sometimes been called into question [10], and some of their items have received criticism for being either too narrowly defined (only applicable to a single situation) or overly broad (too vague or subject to interpretation) [11]. Furthermore, while pain-related avoidance can occur due to fear of injury or reinjury, fear of increased pain, or an actual increase in pain, none of these questionnaires attempt to distinguish between these different cases [12].

In an attempt to address these shortcomings, Neblett et al. (2015) have developed the Fear Avoidance Component Scale (FACS) [9], which includes items from other published FA related measures (TSK, PASS, PCS, FABQ). The FACS also includes items based on the Injustice Experience Questionnaire (IEQ) [13], designed to assess the degree to which chronic pain sufferers feel injustice in relation to their pain. All FACS items were created to assess specific fear avoidance related beliefs and feelings about a person’s painful medical condition, such as cognitive (pain catastrophizing), affective (pain-related fear and anxiety), and behavioral (avoidance) constructs [9]. In addition, six items (15 to 20) were developed to assess the specific types of activities and physical intensity of activities (from low to strenuous) that an individual avoids, and three items were developed to evaluate why the individual is avoiding these activities [9].

The original English version of the FACS has shown acceptable test-retest reliability (r=0.90-0.94) as well as acceptable internal consistency (Cronbach α=0.92) [9]. The FACS has been translated and validated in several different languages. It has shown good psychometric properties in Serbian (test-retest reliability: ICC = 0.93; internal consistency: Cronbach α =0.90) [14], Spanish (convergent validity: r=0.41; internal consistency: Cronbach α=0.90-0.88) [15], Gujarati (test-retest reliability: ICC=0.92; internal consistency: Cronbach α=0.83) [16], Dutch (internal consistency: Cronbach α=0.92; test-retest reliability: ICC=0.92, CI 0.80-0.96) [17] and Turkish (internal consistency: Cronbach α=0.815; test-retest reliability: ICC=0.53-0.97).

There are currently about 321 million French speakers throughout the world. [18] However, a psychometrically validated French version of the FACS has not been made available to date. Clinical settings in French-speaking parts of the world – including France and Canada – could certainly benefit from a French version of this questionnaire. The aim of the current study was to translate the FACS into a common French version, including dialects of France and Canadian French (FACS-Fr/CF), and to assess the psychometric properties of the translated questionnaire – including internal consistency, test-retest reliability, and construct validity.

## 2 Materials and Methods

The study was approved by the ethics committee of the Research Center on Aging du Centre Intégré Universitaire de Santé et de Services Sociaux de l’Estrie – Centre hospitalier universitaire de Sherbrooke (CIUSSS de l’Estrie CHUS) and registered on the ClinicalTrials website (NCT05217017). This study was conducted according to the principles expressed in the Declaration of Helsinki [19].

### 2.1 Cross-cultural translation process

The translation was performed using a six-step process according to the guidelines for the cross-cultural adaptation process written by the American Association of Orthopaedic Surgeons [20]. Initial translation, synthesis, back translation, expert committee, test of the prefinal version and submission of the document to the one of the original developers (RN) were performed. For the initial translation, two independent translators (ST and SW), whose mother tongue was French, translated the scale from English to French. A synthesized version of the two translated questionnaires was completed after discussion with the translator and research team. Two independent translators (BVD and AS), blinded to the original scale and whose native language was English, then translated the synthesized version back to English. An expert committee, comprised of the four translators and the research team, consolidated the prefinal version.

The content validity of the French FACS was pre-tested by 10 healthcare professionals (including physiotherapists, nurses, neuropsychiatrists, and orthopedic surgeons) and 20 individuals (healthy and pain patients), including 10 in Quebec (Canada), and 10 in France. All of them tested the prefinal version by completing the questionnaire and evaluating their understanding of each item. Each person was invited to report any interpretation difficulties and other observations about each test item [20]. Based on participants’ answers, only item 13 « *La douleur causée par mon état de santé est un signal d’alerte indiquant que quelque chose ne va pas du tout chez moi* » (“The pain from my medical condition is a warning signal that something is dangerously wrong with me”) turned into « *La douleur causée par mon état de santé est un signal d’alerte indiquant que quelque chose ne va pas du tout* » (“The pain from my medical condition is a warning signal that something is dangerously wrong”). This modification was made because of the end of the sentence (« *chez moi* »), suggesting a psychiatric disorder connotation in French. For the final step, the methods for obtaining the corrected French version (FACS-Fr/CF) were submitted to author (RN), one of the developers of the original FACS questionnaire (Fig 1). The cross-cultural translation process period ran from March 14, 2022 to March 31, 2022.

**Fig 1.**
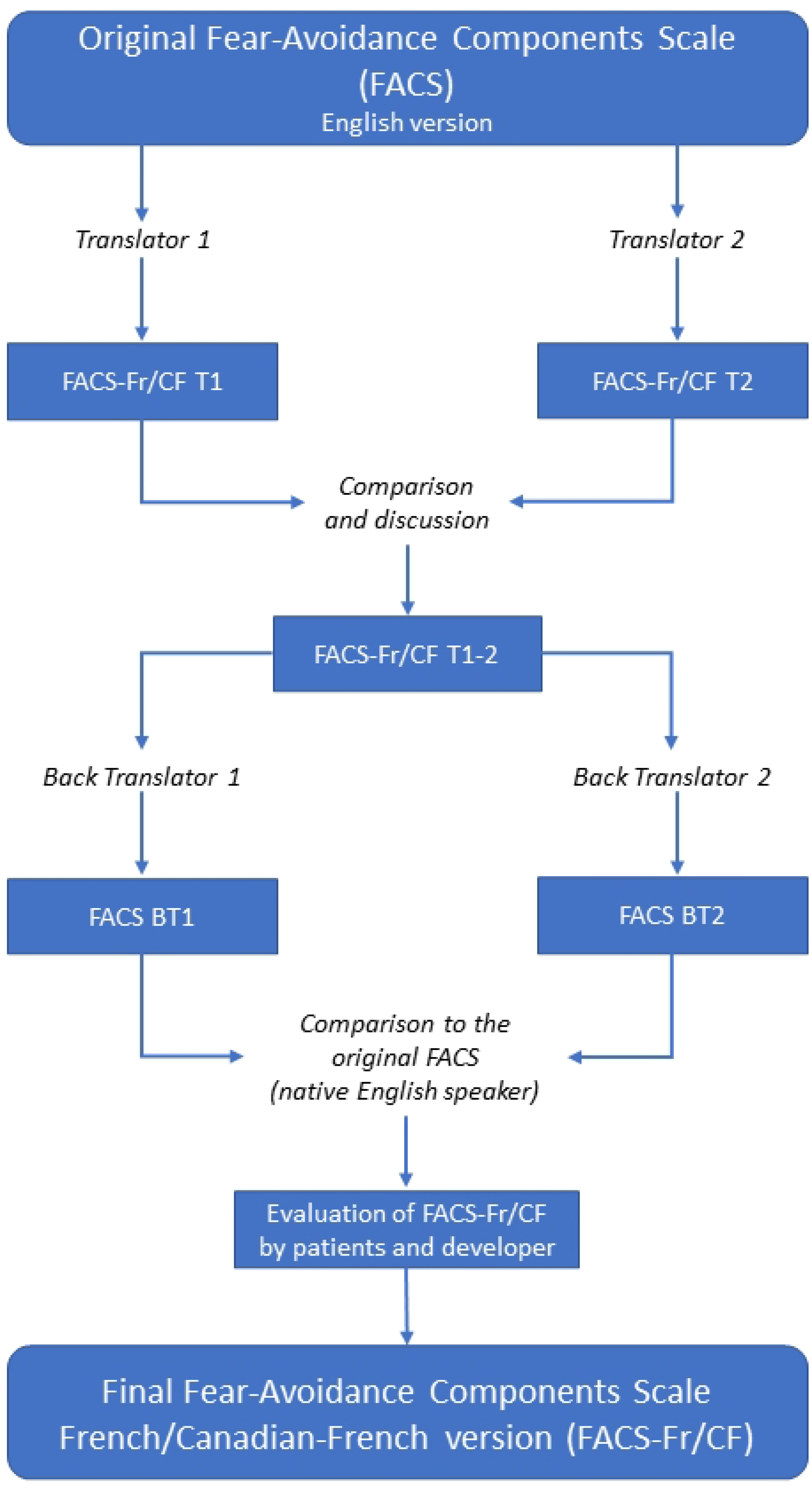
Flowchart of the development of the FACS-Fr/CF.

### 2.2 Psychometric evaluation of the FACS-Fr/CF

After cross-cultural adaptation and translation, the final version of the FACS-Fr/CF questionnaire was administered to a sample of participants suffering from chronic pain to evaluate internal consistency, test-retest reliability, and convergent validity. This validation step was performed according to specialized pain medicine guidelines [21].

#### 2.2.1 Study population

The target population for this study included patients with chronic musculoskeletal pain who spoke French as their first language and referred by a physician for a musculoskeletal condition at the hospital CIUSSS de l’Estrie-CHUS (convenience sampling method). The eligibility criteria were: 1) 18 years of age or older; 2) French as first language; and 3) chronic musculoskeletal pain for at least 3 months. Subjects were excluded if they were unable to consent, read or understand the study requirements (see Fig 2). The inclusion period ran from April 26, 2022 to November 1, 2022.

**Fig 2.**
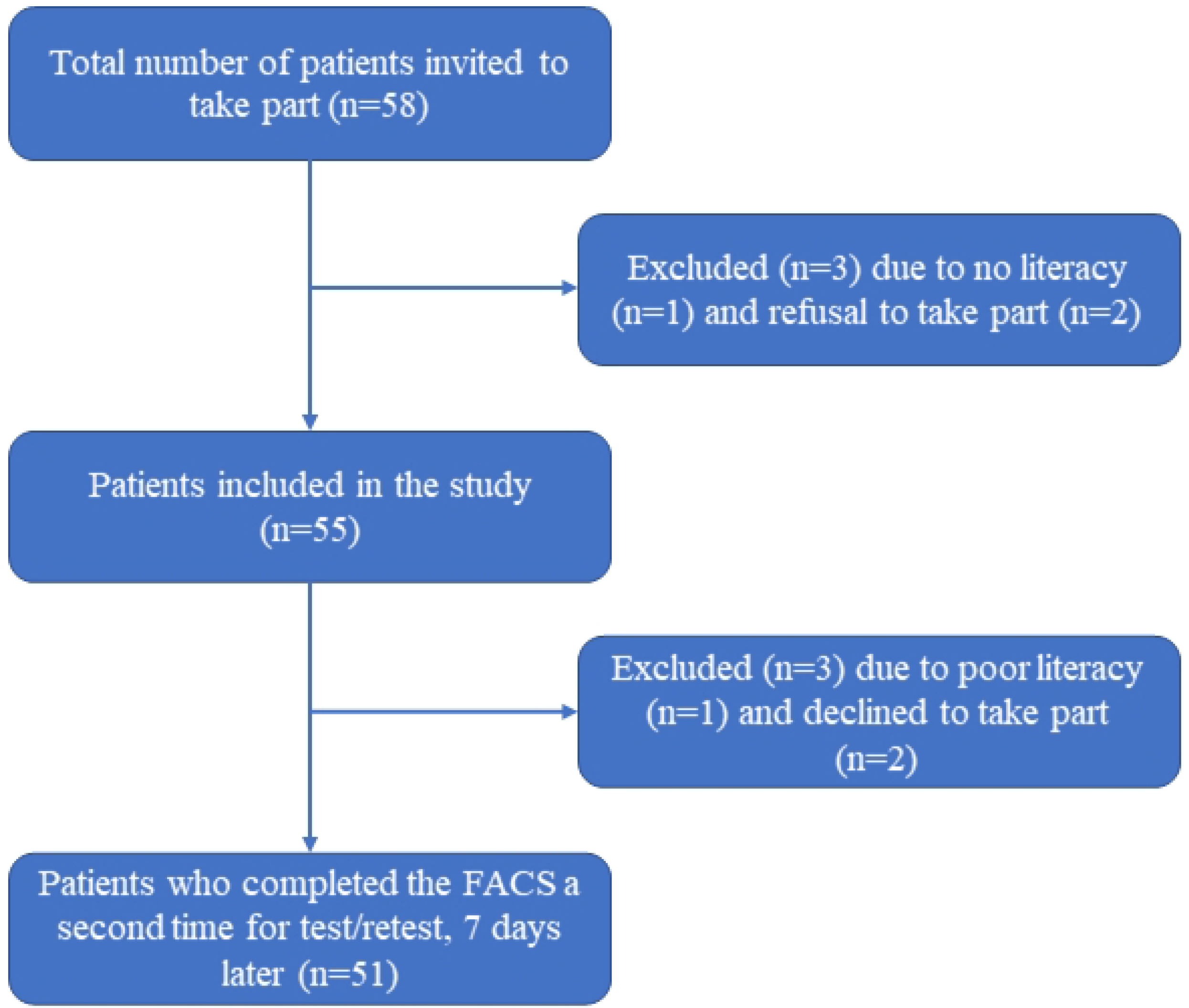
Patient recruitment flowchart.

#### 2.2.2 Sample size

To follow the Consensus-based Standards for the Selection of Health Measurement Instruments recommendations, we recruited 55 participants to meet the objectives of this study (50+10% loss) for the assessment of internal consistency, reliability and validity [22]. This percentage of loss to follow-up was based on the latest research at the CHUS orthopedic service and on a systematic review focused on orthopedic clinical services [23].

#### 2.2.3 Patient-reported outcome measures

The Fear-Avoidance Components Scale (FACS) is a self-reported questionnaire used to comprehensively measure and identify FA in patients with painful medical conditions [24]. It brings together major FA components from earlier FA scales while seeking to address some of their inadequacies within the context of the most recent FA model [11]. This questionnaire consists of 20 items on a Likert scale, ranging from 5 (completely agree) to 0 (completely disagree). The total score, which varies from 0 to 100, is calculated by adding the values of each item. The FACS includes 5 severity levels with increasing severity, based upon quintiles: Subclinical (0 to 20), Mild (21 to 40), Moderate (41 to 60), Severe (61 to 80), and Extreme (81 to 100). Psychometric properties of the FACS show high internal consistency (α = 0.92) and high test-retest reliability (r = 0.90-0.94, P < 0.01) [24]. The Serbian version of FACS found 2 different factors; factor 1 dealt with “general fear avoidance” and included items 1-14, while factor 2 was related to “types of activities that are avoided” and included items 15-20) [14].

The Tampa Scale for Kinesiophobia - Canadian French version (TSK-CF) is a self-reported questionnaire used to assess kinesiophobia. The TSK-CF has demonstrated good psychometric properties (Cronbach’s alpha = 0.71 and ICC > 0.7) [25,26]. This questionnaire consists of 17-items evaluated on a Likert scale, ranging from 1 (strongly disagree) to 4 (strongly agree). The total score, which varies from 17 to 68, is calculated by adding the values of each item. There is no specific threshold to indicate a clinically disabling level of kinesiophobia [27].

The Pain Catastrophizing Scale - Canadian French version (PCS-CF) is a self-reported questionnaire used to measure catastrophic thoughts. The PCS-CF has shown good psychometric properties (Cronbach’s alpha = 0.87 and ICC > 0.7) [28–31]. This questionnaire consists of 13 items, rated on a Likert scale from 0 (not at all) to 4 (always), that can be categorized into three subscales: rumination (being unable to stop thinking about how much it hurts), amplification (exaggerating the threat value of pain sensations), and feelings of helplessness (feeling unable to cope with pain) [32]. The sum of the 13 items’ values yields the final score, which ranges from 0 to 52. It has been suggested that a threshold score of 30 or higher can be used to identify people who have a clinically significant level of pain catastrophizing [33].

The Hospital Anxiety and Depression Scale - French Canadian version (HADS-FC) is a self-reported questionnaire used to measure the symptoms of anxiety and depression [34]. This questionnaire consists of 14 items intended to evaluate the severity of anxiety and depressive symptoms on two different subscales using a 4-point Likert-type scale (ranging between 0 and 3). Higher scores on the total scale indicate greater psychological distress. The internal consistency of the French version is good [35], with the depression subscale having Cronbach’s alpha of 0.78 and the anxiety subscale having a Cronbach’s alpha of 0.81.

#### 2.2.4 Study process and procedures

Participants were recruited during an initial medical consultation visit at the orthopedic clinic of the CIUSSS de l’Estrie – CHUS. After the orthopedist completed the examination and consent to participate was obtained, patients were asked to complete a short sociodemographic questionnaire (including sex, age, physician’s diagnosis, pain duration and academic level) using an online REDCap platform (REDCap 12.4.2 - © 2023 Vanderbilt University) [36]. Patients were then asked to fill out the four questionnaires (FACS-Fr/CF, TSK-CF, PCS-CF, HADS-CF). The questionnaires were completed online (REDCap) or on paper, depending on the participants’ convenience or preference. Then, seven days later, the patients were asked to complete the FACS-Fr/CF a second time, in the same way as the first time (online or paper). This timeframe was short enough to avoid significant clinical fluctuations from first completion and allow an appropriate test-retest evaluation [37].

#### 2.2.5 Statistical analysis

Cronbach’s alpha was used to assess the global internal consistency of the FACS-Fr/CF. We performed a global and confirmatory factors analysis (1 and 2) of the Serbian version. According to Wind *et al*., 2005, α ≥ 0.80, α ≥ 0.70 and α < 0.70 are considered as high, moderate and low, respectively [38].

The reliability was calculated with test-retest intraclass correlation coefficients (ICC) and Pearson correlation coefficients. According to Wind *et al*., 2005, ICC ≥ 0.90, ICC ≥ 0.75 and ICC < 0.75 are considered as high, moderate and low, respectively [38].

Convergent validity was assessed using Pearson’s correlation coefficients, comparing the FACS-Fr/CF with the TSK-CF, the PCS-CF, and the HADS-FC. According to Wind *et al*., 2005, r ≥ 0.60, r ≥ 0.30 and r < 0.30 are considered as high, moderate and low, respectively [38]. Considering the constructs of these different questionnaires, we expected to find stronger correlations with TSK-CF, followed by the PCF-CF and the HADS-FC.

All statistical analyses were performed using SPSS^®^ (version 21); the significance level set at p=0.05.

## 3 Results and discussion

### 3.1 Participants’ characteristics

Demographic information is provided in Table 1. Fifty-five (55) participants took part in the study, including 30 men (54.5%) and 25 women (45.5%). The average age for the total sample was 51.15±16.47 years old. The average pain duration was 65.67±86.80 months. Pain-related medical diagnoses included arthrosis (n=12), tendinopathy (n=9), coxalgia (n=8), ligamentoplasty (n=6), labrum tear (n=5), tendon rupture (n=3), and low back pain (n=2). One subject each, for the remaining 10 patients, were diagnosed with the following: stenosing flexor tenosynovitis, femur elongation, cervical surgery, elbow fracture, ankle pain, complex regional pain syndrome (CRPS), neuropathy of upper limbs, patellofemoral syndrome, Baker’s cyst, crowned dens syndrome. The study ended on November 11, 2022 with the receipt of the last questionnaire.

**Table 1.** Participant characteristics.

| **Participant's characteristics** | **Total sample or mean (% or SD)** |
| --- | --- |
| **Age (years)** | 51.15 (16.47) |
| **Sex** | 30 men (54.5%)<br>25 women (45.5%) |
| **Diagnosis** | Arthrosis (12)<br>Tendinopathy (9)<br>Coxalgia (8)<br>Ligamentoplasty (6)<br>Labrum tear (5)<br>Tendon rupture (3)<br>Low back pain (2)<br>Other (10) |
| **Pain duration (months)** | 65.67 (86.80) |
| **FACS-Fr/CF (at first time)** | 47.35 (22.85) |
| **PCF-CF** | 19.40 (13.81) |
| **TSK-CF** | 40.94 (9.71) |
| **HADS-FC** | 13.49 (8.06) |
| **FACS-Fr/CF (retest 7 days later)** | 45.43 (21.84) |

### 3.2 Internal consistency

The global internal consistency of FACS-Fr/CF calculated by Cronbach’s alpha was high (α=0.94, 95% CI: 0.91-0.96). For factors 1 and 2, the Cronbach’s alpha was high for both (α=0.90, 95% CI: 0.86-0.94 and α=0.90, 95% CI: 0.85-0.94 respectively). The descriptive statistics and internal consistency for the FACS-Fr/CF items are shown in Table 2.

**Table 2.** Internal consistency for the FACS-Fr/CF items.

| FACS-Fr/CF items | Corrected total-item correlation | Cronbach's alpha if item deleted |
| --- | --- | --- |
| 1 | .475 | .934 |
| 2 | .519 | .934 |
| 3 | .643 | .931 |
| 4 | .693 | .930 |
| 5 | .677 | .931 |
| 6 | .451 | .935 |
| 7 | .641 | .931 |
| 8 | .740 | .930 |
| 9 | .708 | .930 |
| 10 | .641 | .931 |
| 11 | .727 | .930 |
| 12 | .299 | .938 |
| 13 | .516 | .934 |
| 14 | .617 | .932 |
| 15 | .634 | .932 |
| 16 | .722 | .930 |
| 17 | .617 | .932 |
| 18 | .826 | .928 |
| 19 | .695 | .930 |
| 20 | .652 | .931 |

### 3.3 Reliability test retest

Test-retest reliability of the FACS-Fr/CF was moderate, close to high (ICC=0.89; 95% CI: 0.82-0.94 and r=0.89; *p*<0.005) [39].

### 3.4 Convergent validity

The convergent validity was assessed with Pearson’s correlation coefficients (unilateral). The FACS-Fr/CF scores were highly correlated with scores on the TSK-CF (r=0.82; *p* < 0.005), PCS-CF (r=0.72; *p* < 0.005) and HADS-FC (r=0.66; *p* < 0.005). All correlation coefficients are shown in Table 3.

**Table 3.**
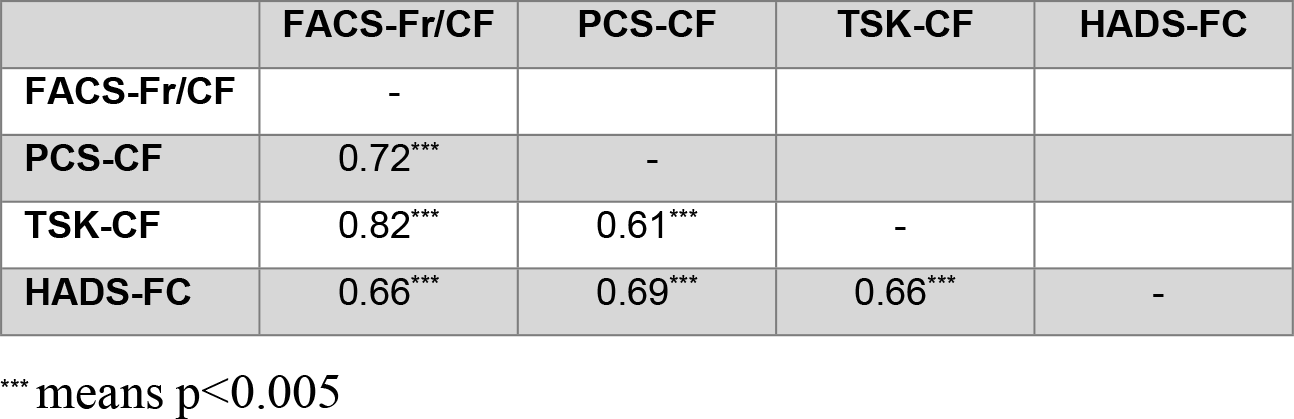
Pearson’s correlations (2-tailed) among PROs.

### 3.5 Discussion

This study aimed to establish and validate a cross cultural adaptation of the FACS questionnaire in French and Canadian French, using the guidelines for questionnaires in pain medicine proposed by Tsang, Royse and Terkawi [21]. Following a forward and backward translation process, and feedback from 10 healthcare professionals and 20 patients, a final version of the FACS-Fr/CF was established. The psychometric properties were then assessed in a sample of 55 chronic pain patients who completed the final FACS-Fr/CF on 2 occasions at a 2-week interval, as well as 3 other questionnaires, which assessed related constructs.

Global internal consistency of FACS-Fr/CF was high (α=0.94,) and comparable to the original FACS (α=0.92) [24]. The internal consistency of the 2 factors identified previously in the Serbian version was also high, despite the relatively small number of items for factor 2. Test-retest reliability of the FACS-Fr/CF was moderate, very close to high. These values are consistent with those obtained for the original version (r = 0.90-0.94, P < 0.01) [24].

For convergent validity, we assessed the relationship between the scores of the FACS-Fr/CF, the TSK-CF, the PCS-CF and the HADS-FC. A previous study with the Spanish and Turkish versions of the FACS found a moderate and strong correlation with the Spanish and Turkish version of the TSK, with a coefficient r=0.39 and r=0.56 respectively [40,41]. In the present study, we observed a high correlation between the FACS-Fr/CF and the TSK-CF (r=0.82). As expected, the correlation was higher with the TSK-CF, compared to the PCS-CF (r=0.72) and the HADS-FC (r=0.66), suggesting that the constructs underlying the FACS and TSK are particularly close [24]. This finding is perhaps not so surprising when we bear in mind that some of the items from the FACS were borrowed directly from the TSK [9].

We also examined the convergent validity of FACS, by assessing its relationship with the PCS. In previous studies, the Spanish version showed a moderate correlation with the PCS (r=0.49 to 0.53) [40,42], and the Serbian and Turkish versions showed a high correlation (r=0.77 and r=0.68 respectively) [14,41]. The high correlation between the FACS-Fr/CF and PCS-CF (r=0.72; *p* < 0.005) was very close to the Serbian and Turkish versions.

FA is frequently associated with anxiety [43] and depression [44]. Though anxiety and depression are related to FA, these constructs are somewhat different [45]. The association between FACS-Fr/CF and HADS-FC scores was therefore expected to be weaker than those observed between the FACS-Fr/CF and TSK-CF, and between the FACS-Fr-CF and PCS-CF.

All in all, the pattern of convergent validity results in the present study is consistent with the a priori assumptions that postulated a higher correlation coefficient between FACS-Fr/CF and TSK-CF scores [46,47], followed by the PCS-CF (with symptoms of helplessness, rumination and magnification being an important component of the FA model [48]) and by the HADS-FC [49].

This study has a number of strengths and limitations. One strength was that the American Association of Orthopedic Surgeons and the Consensus-based Standards for the Selection of Health Measurement Instruments recommendations were followed in the cross-cultural adaptation and validation process. Furthermore, the FACS-Fr/CF was developed as a standardized version, accommodating two distinct French language dialects (France and Canada). As a result, it has potential for broad applicability across diverse French-speaking regions, worldwide. However, it is important to exercise caution when using the instrument in specific contexts, as further validation studies may be necessary to ensure its appropriateness for local linguistic variations and cultural nuances.

An important limitation of this study concerns recruitment, as the 55 patients included in the sample were recruited from a single hospital – a situation that may raise questions about the generalizability of the results. However, it is important to note that this limitation may be partially mitigated by the important variability observed among the pain conditions encompassed within the sample. Nonetheless, extrapolating the results to broader chronic pain populations, outside the specific context of the study, should be done with caution.

## 4 Conclusions

This is the first translation, intercultural adaptation and validation study of the FACS in French version, including dialects of France and Canadian French. The FACS-Fr/CF showed a high global internal consistency and moderate (very close to high) test-retest reliability. The convergent validity of the FACS-Fr/CF was demonstrated by positive correlations with TSK, PCS and HADS. This work provides an important basis for the future use of the FACS-Fr/CF in assessing fear-avoidance beliefs in various French-speaking cultural contexts.

## 5 Supporting information

**S1 File. Fear-avoidance Components Scale, French/Canadian French version (FACS-Fr/CF)**

## Data Availability

Data cannot be shared publicly due to restrictions imposed by the ethics committee, as data sharing was not mentioned in the consent form. Data are available from the Research Center on Aging du Centre Intégré Universitaire de Santé et de Services Sociaux de l’Estrie – Centre hospitalier universitaire de Sherbrooke (CIUSSS de l’Estrie CHUS) Ethics Committee (contact via) for researchers who meet the criteria for access to confidential data.

## Acknowledgments

We would especially like to thank Doctor Stéphane Ricard and the orthopedic service of CIUSSS de l’Estrie-CHUS for recruiting participants, as well as Professor Cynthia Gagnon for her help with the translation process.

